# Extracellular Vesicles as Biomarkers for Steatosis Stages in MASLD Patients: an Algorithmic Approach Using Explainable Artificial Intelligence

**DOI:** 10.1101/2024.12.07.24318644

**Authors:** Eleni Myrto Trifylli, Athanasios Angelakis, Anastasios G. Kriebardis, Nikolaos Papadopoulos, Sotirios P. Fortis, Vasiliki Pantazatou, Ioannis Koskinas, Hariklia Kranidioti, Evangelos Koustas, Panagiotis Sarantis, Spilios Manolakopoulos, Melanie Deutsch

## Abstract

**Background & Aims:** Metabolic dysfunction-associated steatotic liver disease (MASLD), formerly known as NAFLD, is a leading cause of chronic liver disease worldwide. Current diagnostic methods, including liver biopsies, are invasive and have significant limitations, emphasizing the need for non-invasive alternatives. This study aimed to evaluate extracellular vesicles (EV) as biomarkers for diagnosing and staging steatosis in MASLD patients, utilizing machine learning (ML) and explainable artificial intelligence (XAI).

**Methods:** This prospective, single-center cohort study was conducted at the GI-Liver Unit, Hippocration General Hospital, Athens. It included 76 MASLD patients with ultrasound-confirmed steatosis and at least one cardiometabolic risk factor. Patients underwent transient elastography for steatosis and fibrosis staging and blood sampling for EV analysis using nanoparticle tracking. Twenty machine learning models were developed. Six to distinguish non-steatosis (S0) from steatosis (S1-S3), and fourteen to identify severe steatosis (S3). Models incorporated EV measurements (size and concentration), anthropomorphic and clinical features, with performance evaluated using AUROC and SHAP-based interpretability methods.

**Results:** The CB-C1a model achieved, on average on 10 random splits of 5-fold cross validation (5CV) of the train set, an AUROC of 0.71/0.86 (train/test) for distinguishing S0 from S1-S3 steatosis stages, relying on EV alone. The CB-C2h-21 model identified severe steatosis (S3), on average on 10 random splits of 3-fold cross validation (3CV) of the train set, with an AUROC of 0.81/1.00 (train/test), demonstrating superior performance when combining EV with anthropomorphic and clinical features such as diabetes and advanced fibrosis. Key EV features, including mean size and concentration, were identified as important predictors. SHAP analysis highlighted complex non-linear relationships between features and steatosis staging.

**Conclusions:** EV are promising non-invasive biomarkers for diagnosing and staging MASLD. The integration of ML-enhanced EV analysis with clinical features offers a scalable, patient-friendly alternative to invasive liver biopsies, advancing precision in MASLD management. Further research should refine these methods for broader clinical application.

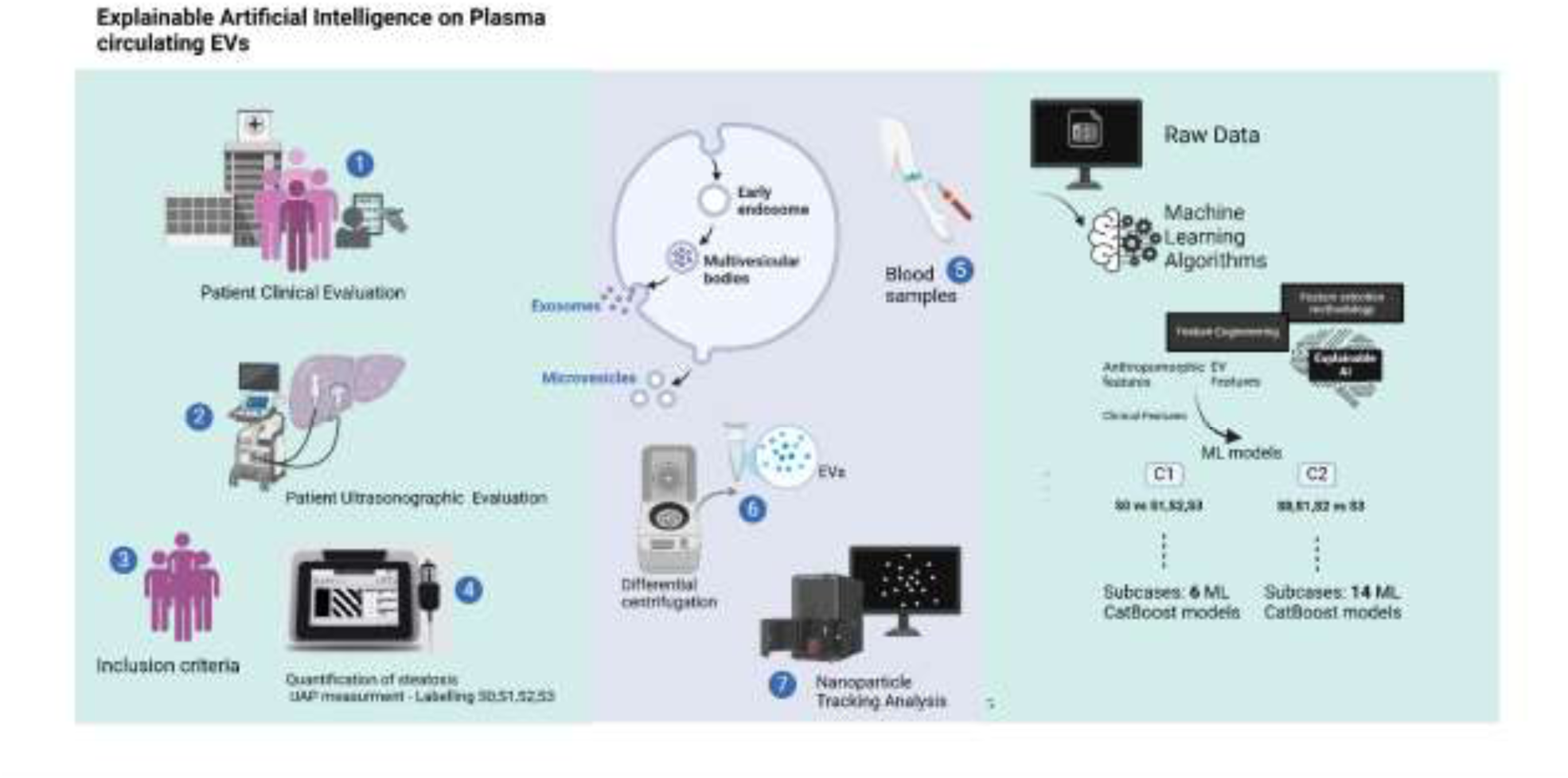

## 1. Introduction

MASLD is one of the most frequently diagnosed and continuously increasing chronic liver diseases, particularly in the Western world [1]. It is characterized by significant morbidity and mortality, especially in advanced stages, and is currently the leading cause of liver transplantation in the US [2]. MASLD includes hepatic steatosis which may progress to metabolic dysfunction-associated steatohepatitis (MASH), fibrosis, cirrhosis, and hepatocellular carcinoma. The main challenges in everyday clinical practice include firstly the diagnosis of MASLD, secondly the diagnosis of MASH and the determination of the steatosis and fibrosis extent [3]. Although the extend of liver fibrosis is the strongest predictor of end stage liver disease, the presence and the degree of steatosis are also important factors that should be accessed since they are closely implicated in the development of hepatic and extrahepatic outcomes. More particularly the two most common causes of mortality in those patients are cardiovascular outcomes and extrahepatic malignancy, while the dissemination of different primary cancer in hepatic parenchyma is closely correlated with the degree of steatosis (HR of 1.34) based on EASL–EASD–EASO Clinical Practice Guidelines, whereas the least common cause is interestingly due liver-related outcomes [4]. Interestingly, MASLD patients have a higher risk of developing extrahepatic malignancies, than obese patients, implying its significant implication in obesity-mediated cancers [4, 5].

Liver ultrasound is the most commonly used diagnostic method, especially for cirrhosis identification. However, it is an operator-dependent diagnostic method and it is considered insufficient for the evaluation of low-grade steatosis or fibrosis [6]. Liver biopsy remains the gold standard for the definitive diagnosis of MASH, and fibrosis evaluation, even at the early disease stages. However, its invasive nature, high cost, and variability in diagnostic accuracy limit its widespread use [7, 8]. Liver stiffness measurement (LSM) with transient elastography (TE) and attenuation parameters are reliable approaches for identifying liver fibrosis and steatosis respectively. However, there is an emerging need of developing new and easily-performed noninvasive tests (NITs) or combinations of tests in order to assess liver or non-liver prognosis in MASLD patients. [9].

In this context, extracellular vesicles (EVs) have emerged as probable diagnostic biomarkers and could be an interesting alternative [10, 11]. EV’s are double-membraned nanoparticles, secreted by multiple cells, that carry several molecules such as DNA, RNA, proteins, lipids, and autophagosomes [10]. EVs significantly contribute to intercellular communication by altering the recipient cells’ functional and biological status [11]. Lipotoxic hepatocytes are a major source of EVs in the systemic circulation. During lipotoxicity, increased lipid storage in liver parenchyma induces the release of EVs, which play a pivotal role in MASLD progression by promoting inflammatory reactions, recruiting macrophages and monocytes, and activating hepatic-stellate and liver sinusoidal endothelial cells, enhancing fibrosis and angiogenesis [10–13]. It is important to underline the crucial role of EVs within the framework of steatotic liver that can induce the development of extrahepatic malignancies and secondary hepatic lesions.

Alterations in EV quantity and quality are closely implicated in MASLD pathogenesis and could facilitate diagnosis, patient stratification, and prognosis [14]. Moreover, it has been demonstrated in animal models (murine) that the EV levels are closely related to the EV circulating levels, especially in cases of MASH compared to healthy controls that have been strongly related to increased angiogenesis and advanced fibrosis [14, 15]. Additionally, it was demonstrated in another study that EVs were increased in pre-cirrhotic patients and were even more abundant in cirrhotic patients, in comparison to patients with advanced MASH and healthy individuals [16]. It has been also demonstrated in the same study by Povero et al. [16], that liver constitutes a significant source of EV in blood circulation. On the top of that, another study has reported a depletion of small-sized EVs after bariatric surgery and the subsequent weight loss [17]. Moreover, it was shown that plasma EVs especially the hepatocyte-derived ones, are notably increased in the case of MASLD patients, in comparison to post-weight loss patients, which implies the potential use of circulating EVs of hepatocyte origin as a diagnostic and prognostic biomarker in a point-of case manner [17]. The patient’s response to surgical intervention (bariatric surgery) or other lifestyle modifications, hints their potential utilization also as monitoring tools for treatment response.

Additionally, several conditions can increase their levels and size such as in cirrhosis and YPE 2 Diabetes Mellitus (T2DM). More particularly, T2DM can alter the levels and the size of the detected plasma EVs [18]. More specifically, it was demonstrated in several studies the presence of higher levels of microvesicles in patients with T2DM, compared to euglycemic individuals [19]. A meta-analysis concluded that the levels of circulating large EVs (microvesicles) were indeed higher in T2DM patients, especially the ones that were derived from monocytes, endothelial cells, and platelets, in contrast to the levels that originated by leucocytes, which have been proven no statistically elevated and inversely correlated with the grade of fibrotic injury [20]. On top of that, there is a close relation between MASLD and LSM progression (≥20% increase in LSM values) in T2DM, which constitutes a statistically significant independent predictor for LSM increase based on the recent study by Huang, Daniel Q. et al. [21].

This study aimed on the assessment of the severity of steatosis, as well as the presence of steatosis, in MASLD patients using a combination of two parameters: circulating levels of plasma EV and the ultrasound attenuation parameter (UAP) measured by TE [22, 23].

Additionally, we employed data science (DS), ML [24] and XAI [25] methods to understand the non-linear relationships between features and predict severe and presence of steatosis in MASLD patients.

## 2 Materials and Methods

### 2.1 Patients

This study is a prospective single-center study GI-Liver Unit of the Second Academic Department of Internal Medicine Hippocration General Hospital University of Athens), which is composed of 76 consecutive patients with ultrasound findings of steatosis and at least one cardiometabolic risk factor as it was suggested on multi-society Delphi consensus on new nomenclature and no other causes of steatosis. More particularly, the cardiometabolic risk factors included, a BMI≥ 25 kg/m2 or increased waist circumference for men (>80cm) and women (>94 cm), low HDL levels for males <40mg/dl and for females <50mg/dl, high triglycerides (>150 mg/dl) or the intake of treatment for dyslipidemia, hypertension (≥130/80 mmHg) or the intake of antihypertensive drug regime, as well as the presence high levels of fasting glucose, or high 2h-postprandial glucose levels or diagnosed diabetes mellitus or high levels of glycosylated hemoglobin or the intake of anti-diabetic treatment [26]. Patients with alcohol abuse (≥2 drinks for females and ≥ 3 drinks for males per day, with 1 drink being equal to 10g of alcohol), inflammatory or autoimmune diseases, other chronic viral or metabolic liver diseases, hepatocellular cancer, or non-hepatic malignancy have been excluded [26]. After acquiring the informed consent of all the participants in this study, we collected demographic data, patient’s history and routine blood tests as well as we performed transient elastography (TE) by an experienced operator. The evaluation was made via the “iLivTouch” FT100 device by Wuxi Hisky Medical Technologies Co., Ltd. (Hisky Med), China. This device performs LSM using TE based on controlled low frequency shear wave synchronously with UAP using one universal probe applicable to patients of different body sizes [24, 25]. Steatosis and fibrosis were assessed with the evaluation of the UAP and the LSM, respectively, after the performance of ≥10 valid measurements, with a ratio between the successful to the total number of measurements ≥ 60%, as well as the ratio between the interquartile range (IQR) to the LSM being ≤ 30% based on the guidelines of World Federation for Ultrasound in Medicine and Biology (WFUMB) [27]. The thresholds for steatosis based on the estimation of UAP value were: ≥S3 296dB/dl, ≥S2 269dB/dl, and ≥S1 244dB/dl [10]. However, we further categorized the patients based on the presence of severe steatosis (≥S3 296dB/dl) , S0-S2 (< 296 dB/dl) and S0 < 244 dB/dl [23]. In Table 1 we provide patient characteristics and in Table 2 the percentages of T2DM and advanced fibrosis in different stages of steatosis.

**Table 1.**
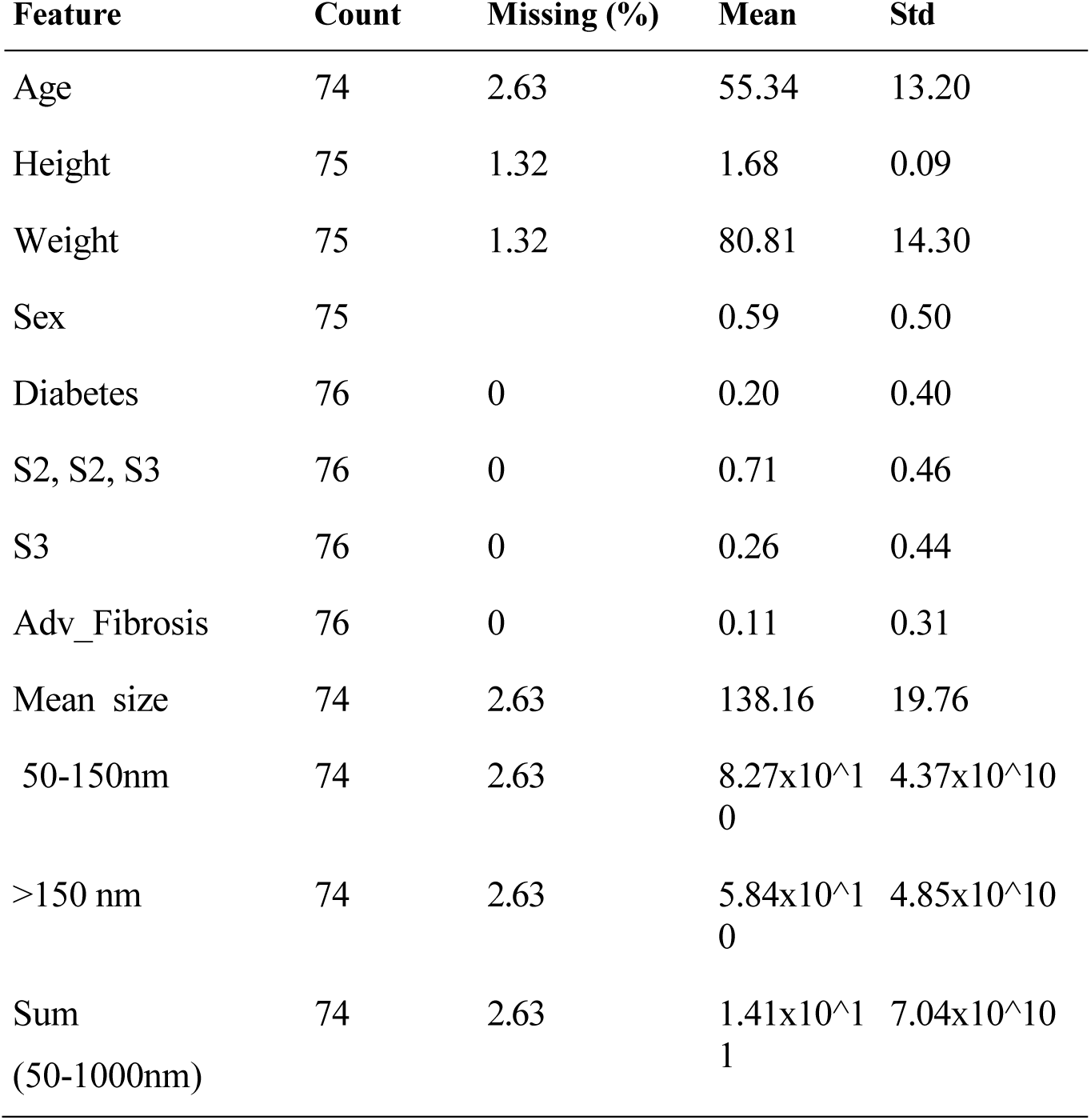
Patient Characteristics.

**Table 2.**
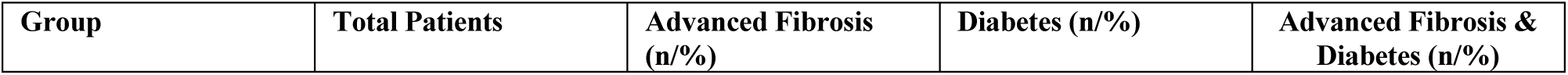
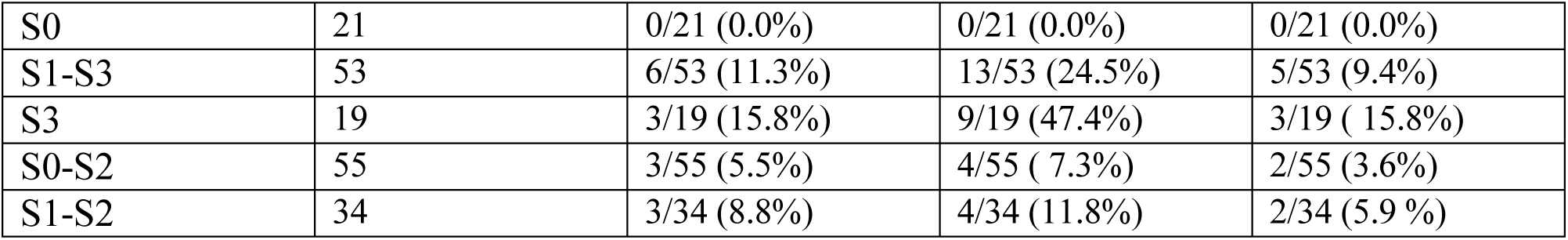
Percentages of advanced fibrosis and diabetes in the groups of S0, S1-S3, S3, S0-S2 and S1-S2.

### 2.2 Pre-analytical protocol

The protocol of pre-analysis was the same for each blood sample, including the time of collection, the blood processing interval (BPI), the handling, and the processing to minimize the variability of EV analysis. We chose the collection tubes with citrate for EV analysis, while the collection of the samples followed the Declaration of Helsinki (DoH) after the patients had given written informed consent. Blood drawing was performed at the same time of day, while the patients were fasting before the procedure. Blood samples were collected in a tube with Citrate (Vacuette sodium citrate 3.2%, volume 3,5 mL, Greiner Bio-One) [27], the time interval for blood drawing was approximately five minutes and the tube was turned vertically and back 10 times exactly after their collection. All the samples were transported to the lab after the collection in special conditions to avoid deterioration with a BPI of 1 hour. These samples were initially centrifuged for 20 min at 3000 x g at room temperature to deplete cell debris and the collected supernatant was immediately stored at −80°C, until the following processing, including differential centrifugation [28].

### 2.3 Nanoparticle Tracking Analysis (NTA)

Nanoparticle Tracking Analysis (NTA) was conducted using the NanoSight NS300 instrument (Malvern Instruments, Amesbury, UK). Plasma samples were diluted in particle-free PBS (0.02 μm filtered, Cytiva Whatman, UK) for NTA by which we acquired data about their total concentration levels, size (between 50-1000nm), and distribution in the sample [29]. All the measurements were performed under constant flow and temperature, while autofocus was adjusted to ensure clarity by avoiding indistinct particles. The type of camera and laser were sCMOS and green, respectively. More particularly, five 30-second videos were recorded per measurement under the following conditions: cell temperature at 25°C and syringe speed at 100 μL/second. During the measurements, there was no evidence of vibration, which can alter the sizing of particles. Videos were analyzed using NanoSight NTA 3.4 build 3.4.4 software (Malvern, 2020) in script control mode, comprising 1,500 frames per sample. Each sample was measured five times, and the size distribution data were averaged. Additionally, we evaluated the possible inter-day variability of the method, by performing NTA for each sample on different days, which did not show any significant difference between the values. Lastly, we obtained data regarding the mean size of vesicles for each patient, the amount of vesicles sized between 50 to 150nm or >150nm and the sum of vesicles between 50 to 1000nm.

### 2.4 Data Science

Based on the bibliographic data regarding the alterations in EV quantity, size, and quality in MASLD pathogenesis, we utilized ML algorithms and feature engineering (FE) [30], techniques to understand non-linear relationships between features. FE boosts ML performance by creating more features, and feature selection methods identify the most important ones [30]. We applied the CatBoost algorithm [31–34], FE techniques, XAI methods, and feature selection (FS) methodologies [30, 35–38] to create a data science pipeline for predicting non-severe or severe steatosis in MASLD patients in two main cases: Case 1 (C1) and Case 2 (C2), respectively. More specifically, we primarily used UAP cut-off values to label patients between severe and non-severe steatosis and quantified their circulating plasma EV levels to show associations and potential diagnostic performance for C1: S0, S1, S2 vs. S3 and for C2: S0 vs. S1, S2, S3.

To further explore the relationships between features and steatosis stages, we conducted a comprehensive correlation analysis [39] using both parametric and non-parametric tests, including Pearson, Kendall, Spearman, and Point-Biserial correlations [40–43]. This analysis was performed separately for both C1 (S0 vs. S1, S2, S3) and C2 (S0, S1, S2 vs. S3) cases. Correlations were evaluated against a moderate threshold of 0.6, chosen to identify meaningful yet not overly stringent associations. This approach enabled us to assess both linear and rank-based relationships, providing a robust understanding of the interactions between features and steatosis stages.

We used the following EV features: ‘mean’, ‘50-150nm’, ‘>150’, and ‘Sum’, anthropomorphic ones: ‘Sex’, ‘Age’, ‘Height’, ’Weight’, ‘BMI’ and clinical: advanced fibrosis ‘Adv_Fibrosis’, and ‘Diabetes’. Concretely, the ‘mean’ represents the mean size of vesicles for each patient, ‘50-150nm’ the amount of vesicles sized between 50 to 150nm , “>150” the vesicles sized over 150nm and the “sum” that represents the sum of vesicles between 50 to 1000nm. We tuned twenty CatBoost ML models. Additionally, we used XAI and FS [35–38] to identify the most important features. For tuning and to understand the robustness of our models, we used non-stratified k-fold cross validation 5CV, 3CV for C1 and C2 respectively, and a test-set to measure the performance of our ML models. Moreover, since the dataset’s volume was relatively low we used ten times k-fold cross-validation, splitting with different random seeds, to eliminate possible randomness splitting effect [30].

#### 2.4.1 Cases Definition

We investigated the role of EV as biomarkers in two cases. Namely, in C1 we defined the binary classification problem of distinguishing non-steatosis vs steatosis S0 vs. S1, S2, S3. In C2 we defined the binary classification problem of identifying an individual’s severe steatosis stage S0, S1, S2 vs. S3.

C1: We created six sub-cases:

- C1a: Using EV features (mean, 50-150nm, >150, Sum).
- C1b: Applying FE on C1a.
- C1c: Combining EV with anthropomorphic features (Sex, Age, Height, Weight, BMI).
- C1d: Applying FE on C1c.
- C1e: Combining EV features with the presence of advanced fibrosis and diabetes.
- C1f: Applying FE on C1e.

C2: We created eight sub-cases:

- C2a: Using only EV features.
- C2b: Applying FE on C2a.
- C2c: Combining EV features with anthropomorphic features.
- C2d: Applying FE on C2c.
- C2e: Combining EV features with the presence of advanced fibrosis and diabetes.
- C2f: Applying FE on C2e.
- C2g: Combining EV features with anthropomorphic features and the presence of advanced fibrosis and diabetes

C2h: Applying FE on C2g.

#### 2.4.2 Data Preprocessing

For the C2 we used the cut-off UAP value of >= 296 to label the ‘target’ feature, which was severe steatosis (S3) and not severe steatosis (S0, S1, S2). For the C1 we used the cut-off UAP value of >= 244a. We dropped data instances with null values and no rnomalization/scaling applied.

#### 2.4.3 Datasets Creation

For both cases, we split the initial dataset of 74 or 72 data instances, depending on the subcase, to 80% as train and 20% as a test set with a fixed random seed for reproducibility. The percentage of the ‘target’ was similar in the train and test sets in both cases so that the ML models we tuned would be fair. A patient who belonged to the train datasets did not belong to the test sets so there would be no data/information leakage [39]. Furthermore, our approach to random split to train and test set is aligned with the guidelines from the DELFI TRIPOD-AI statement [40] so that our ML models would be applied to the examination center where the data have been retrieved from. The feature selection methodology [6a-9a] uses both datasets but the inductive learning [41] approach of it makes sure there is no information/data leakage [39]. We had two approaches for the examination of the role of EV regarding their predictability and associativity, due to the following reasons: (i) lack of bibliographic data about the assessment of EV role and their predictability on MASLD using XAI and ML models and (ii) the relatively low volume of data instances of the dataset (number of patients). In FE sub-cases we divided the EVs with 10^9, we raised the numeric features to 11^th^ power and we calculated their square root, and finally we calculated the pair-wise products for all features.

#### 2.4.4 Feature selection methodology

Applying the FE methodology [35–38] on the sub-cases C2d, C2f and C2h, lead us to five, eleven and twenty-one respectively, most important features. Analyzing these features using SHAP [42] and CatBoost’s feature importance, provided us with insights regarding the features in C2 cases.

#### 2.4.5 Models Evaluation

All ML models tuned using k-fold cross validation (kCV) on the train dataset for each case, and have been applied to the corresponding per case test set for validation. We used 5 and 3 folds for the C1 and C2 respectively. Each model could correspond to potentially a new method for classifying in an automatic way individuals that belong to each of the classes for each case. Since the data volume was relatively small, after the tuning we applied ten splits with different random seeds for the kCV to eliminate the impact of randomness. We tuned the iterators, the learning rate, the depth and the weights from CatBoost. For all the other hyperparameters we used the default values.

#### 2.4.6 Algorithm

A deterministic algorithm to create two predictive ML models for the cases C1 and C2, and to identify the most important features and their associations.

- Input: VE, anthropomorphic, clinical features and the UPA values of the 76 patients described in 2.1.

- Output: a. The best predictive model for the case C1 and the best one for the case C2.

b. The most important features for each model and their associations to the outcome

**1.** Initialize Data and Libraries:

- Import necessary libraries (CatBoost, SHAP, etc.).
- Load the dataset.
**2.** Data Preprocessing:

- Drop data instances with null values.
- Label the target feature for C1 (S0 vs. S1, S2, S3) and C2 (S0, S1, S2 vs. S3) using the cut-off UAP values (≥ 244a for C1 and ≥ 296 for C2).
**3.** Dataset Splitting:

- Split the dataset into train (80%) and test (20%) sets using a fixed random seed to ensure reproducibility.
- Ensure similar percentages of ‘target’ in both train and test sets.
**4.** Features’ Combinations:

> For each case (C1 and C2), create at least four sub-cases that correspond to:
>
> - Use only EV features.
> - Combine EV with anthropomorphic features.
> - Combine EV with clinical features.
> - Combine EV with anthropomorphic and clinical features.
**5.** Feature Engineering

- For numeric features:

- Divide by 10^9.
- Raise to the 11th power.
- Calculate square roots.
- For all features compute the pair-wise products
**6.** Model Training:

- Use CatBoost model for training.
- Perform k-fold cross-validation (kCV) for tuning hyperparameters (iterators, learning rate, depth, weights).
**7.** Model Evaluation:

- Evaluate the ML models validating them on the test set.
**8.** Feature Selection Methodology:

- Apply FS
**9.** Identify the best model for each case:

- From the statistics on the kCV and the performance on the test set identify the best model for each case.
**10.** Identification of Important Features:

- After feature selection, identify the most important features using SHAP.
**11.** Identify the most important combination of family of features: Conclude about the contribution of each one of the families of features (EV, anthropomorphic, clinical) for C1 and C2

## 3 Results

### 3.1 C1

In C1, the training set consisted of 59 individuals, and the test set included 15. Six CatBoost models were developed, tuning hyperparameters such as iterations, learning rate, depth, and class weights. Performance metrics, including sensitivity, specificity, ROC-AUC, and F1 scores, are summarized in Table 3. The CB-C1a model emerged as the best-performing configuration, achieving an AUROC of 0.70 (±0.15) and an F1 score of 0.73 (±0.14). The model demonstrated balanced performance with a sensitivity of 0.66 (±0.17) and specificity of 0.74 (±0.14).

**Table 3.** ML models of the C1: the name of the model, the number of features for each case, the values of their tuned hyper-parameters, the classes’ weights, the specificity, sensitivity, ROC-AUC, F1 scores, their mean and standard deviation as well as the 95% confidence interval for the 5CV on the train set and on the test set.

| Model | Feats | Iters | LR | Depth | W0 | W1 | Dataset | Spec. (+/-std) 95%CI | Sens. (+/-std) 95%CI | AUROC (+/-std) 95%CI | F1 (+/-std) 95%CI |
| --- | --- | --- | --- | --- | --- | --- | --- | --- | --- | --- | --- |
| CB-C1a | 4 | 21 | 0.20 | 13 | 0.68 | 0.32 | SCV | 0.74 (+/-0.14) [0.60, 1.00] | 0.66 (+/-0.17) [0.43, 0.89] | 0.70 (+/-0.10) [0.51, 0.78] | 0.73 (+/-0.14) [0.50, 0.89] |
| CB-C1a | 4 | 21 | 0.20 | 13 | 0.68 | 0.32 | Test | 0.83 | 0.89 | 0.86 | 0.89 |
| CB-C1b | 54 | 20 | 0.60 | 3 | 0.68 | 0.32 | SCV | 0.60 (+/-0.33) [0.00, 1.00] | 0.80 (+/-0.07) [0.71, 0.89] | 0.70 (+/-0.15) [0.44, 0.88] | 0.81 (+/-0.06) [0.71, 0.88] |
| CB-C1b | 54 | 20 | 0.60 | 3 | 0.68 | 0.32 | Test | 0.67 | 0.89 | 0.78 | 0.84 |
| CB-C1c | 9 | 10 | 0.10 | 8 | 0.68 | 0.32 | SCV | 0.60 (+/-0.23) [0.29, 0.80] | 0.67 (+/-0.24) [0.47, 1.00] | 0.63 (+/-0.01) [0.63, 0.63] | 0.69 (+/-0.10) [0.61, 0.83] |
| CB-C1c | 9 | 10 | 0.10 | 8 | 0.68 | 0.32 | Test | 0.67 | 0.89 | 0.78 | 0.84 |
| CB-C1d | 125 | 3 | 0.1 | 5 | 0.68 | 0.32 | 10CV | 0.56 (+/-0.26) [0.25, 1.00] | 0.71 (+/-0.19) [0.50, 1.00] | 0.64 (+/-0.18) [0.44, 0.94] | 0.72 (+/-0.13) [0.57, 0.93] |
| CB-C1d | 125 | 3 | 0.1 | 5 | 0.68 | 0.32 | Test | 0.83 | 0.89 | 0.86 | 0.89 |
| CB-C1e | 6 | 6 | 0.70 | 8 | 0.68 | 0.32 | SCV | 0.69 (+/-0.21) [0.50, 1.00] | 0.69 (+/-0.29) [0.20, 1.00] | 0.69 (+/-0.19) [0.35, 0.93] | 0.63 (+/-0.22) [0.31, 0.92] |
| CB-C1e | 6 | 6 | 0.70 | 8 | 0.68 | 0.32 | Test | 0.75 | 1.00 | 0.93 | 0.92 |
| CB-C1f | 11 | 2 | 0.60 | 4 | 0.65 | 0.35 | SCV | 0.63 (+/-0.34) [0.00, 1.00] | 0.75 (+/-0.16) [0.50, 1.00] | 0.69 (+/-0.20) [0.39, 0.99] | 0.78 (+/-0.12) [0.62, 0.90] |
| CB-C1f | 11 | 2 | 0.60 | 4 | 0.65 | 0.35 | Test | 1.00 | 0.80 | 0.90 | 0.89 |

We conducted correlation analysis on C1. Across all correlation tests (Pearson, Kendall, Point-Biserial, and Spearman), Figure certain features, such as transformed BMI and diabetes markers, exhibit moderate associations with steatosis stages. However, none of the correlations surpass the 0.6 threshold, indicating that no feature demonstrates a moderately strong or strong relationship with steatosis severity in this comparison. This suggests that the relationships between individual features and the steatosis stages are relatively weak and potentially complex.

The violin plots in the top row of Figure 3 depict the distributions of selected features (>150, >150 nm particle count, mean, and sum). These plots do not reveal any distinguishable patterns or separation between the groups, indicating that these features, when analyzed individually and linearly, fail to provide insight into the differences between steatosis stages. This lack of separation highlights the challenge of identifying meaningful patterns using traditional visualization techniques, particularly for multifactorial diseases like MASLD. Conversely, the SHAP scatter plots in the bottom row, derived from XAI, reveal non-linear patterns in the associations between these features and the target outcome. The scatter plots show that even in cases where traditional statistical methods and visualizations fall short, the XAI approach uncovers nuanced relationships. For example, SHAP values illustrate the varying impact of features like >150, >150 nm particle count, mean, and sum on the classification of steatosis stages, demonstrating their combined importance in the predictive model.

**Figure 1.**
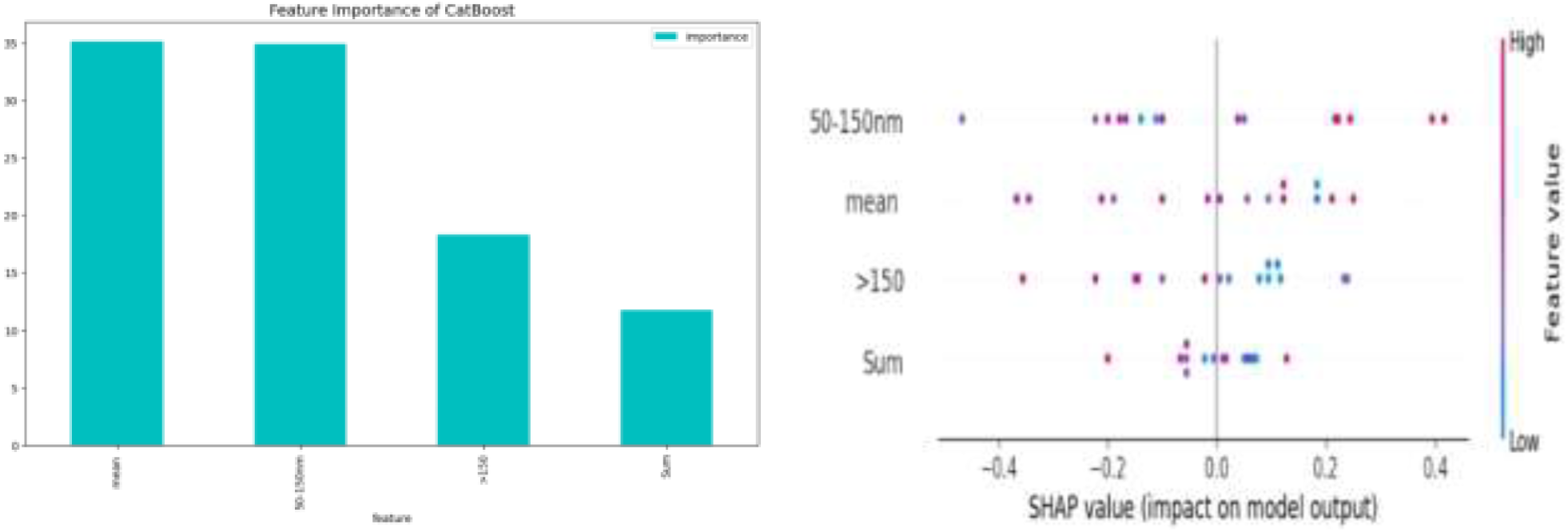
Plots of feature importance on the predictability of the ML model CB-C1a and on SHAP on the train set (ROC-AUC train: 0.70, test: 0.86).

**Figure 2.**
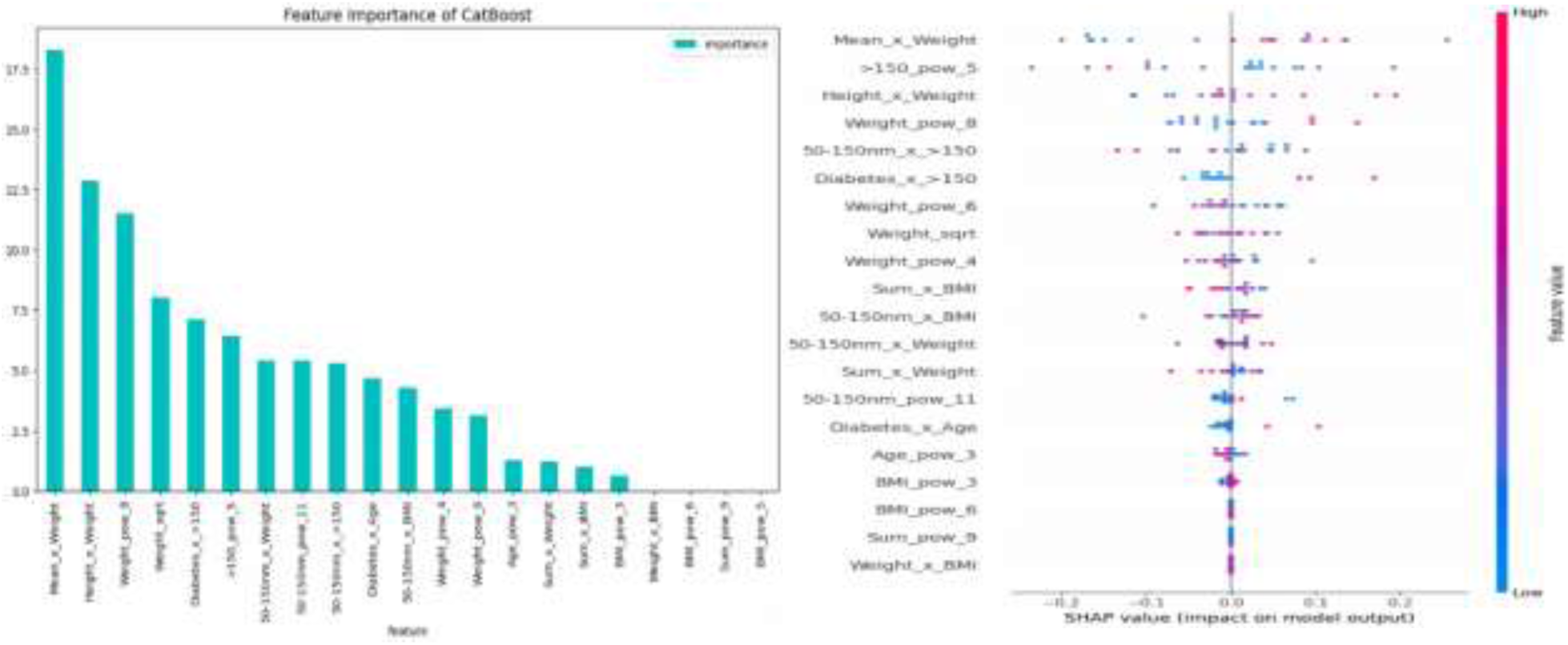
Plots of feature importance on the predictability of the ML model CB-C2h-21 and on SHAP on the train set (ROC-AUC train: 0.89, test: 1.00)

**Figure 3.**
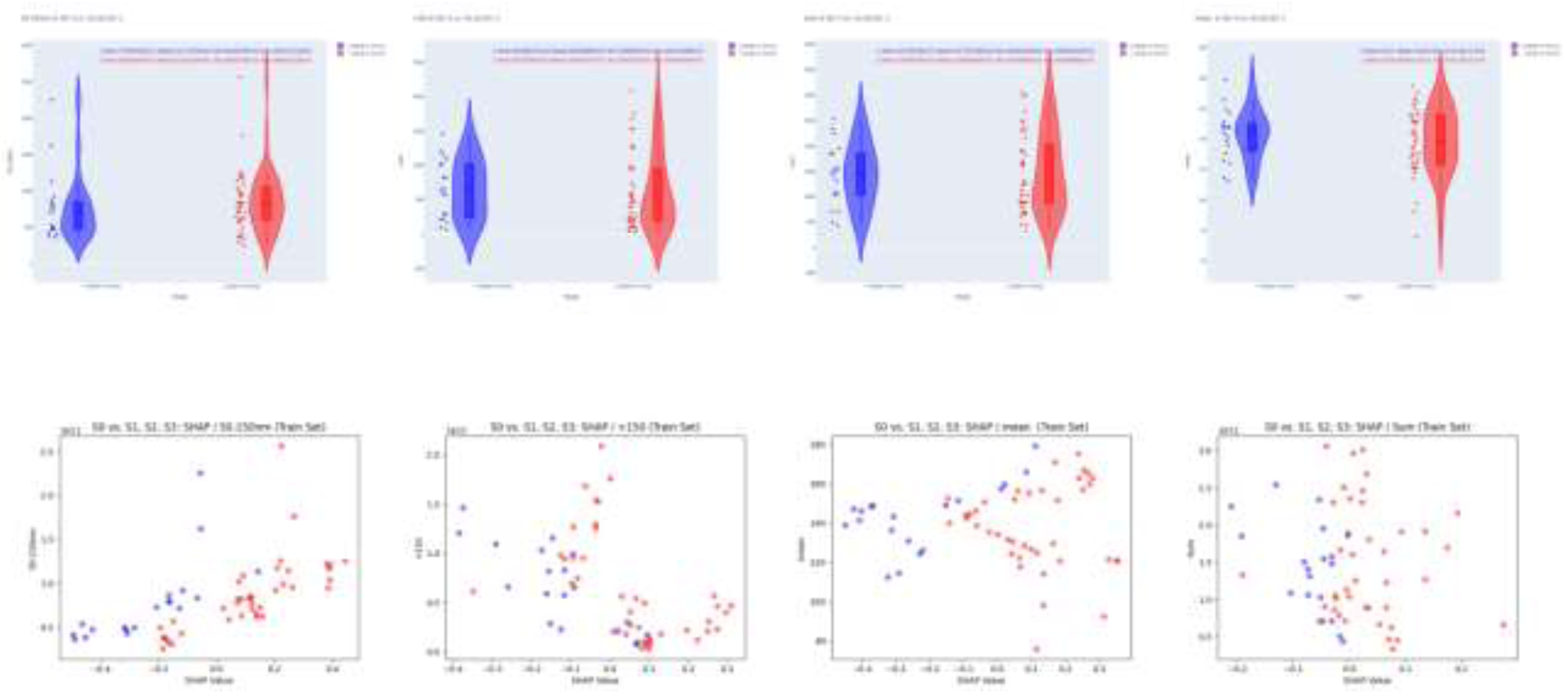
Plots of EVs distribution above and SHAP scatter plots for the train set of the ML CB-C1a model.

The addition of anthropomorphic and clinical features, such as advanced fibrosis and diabetes, or the application of FE, did not improve the model’s performance. In the S1-S3 group, advanced fibrosis was present in only 6/53 cases, diabetes in 13/53, and both conditions in 5/53 (Table 2), supporting the sufficiency of EV for distinguishing S0 from steatosis stages. Feature importance and SHAP analyses, presented in Figure 1, respectively, identified "Mean," "50-150nm," ">150nm," and "Sum" as the most influential predictors, with "Mean" ranking highest. The SHAP scatter plots (Figure 3). highlighted the non-linear relationships between these features and model predictions, showcasing the necessity of advanced ML techniques to capture such complexity.

### 3.2 Case 2: S0, S1, S2 vs. S3 Steatosis

In Case 2, the training set included 58 individuals, and the test set consisted of 13. Fourteen CatBoost models were tuned, and their results are presented in Table 4. The CB-C2h-21 model demonstrated superior performance, with an AUROC of 0.89 (±0.03) on the training set (3CV) and a perfect AUROC of 1.00 on the test set. Sensitivity and specificity values of 0.92 (±0.12) and 0.87 (±0.10), respectively, further underscored its reliability in distinguishing mild-to-moderate steatosis (S0-S2) from severe steatosis (S3).

**Table 4.**
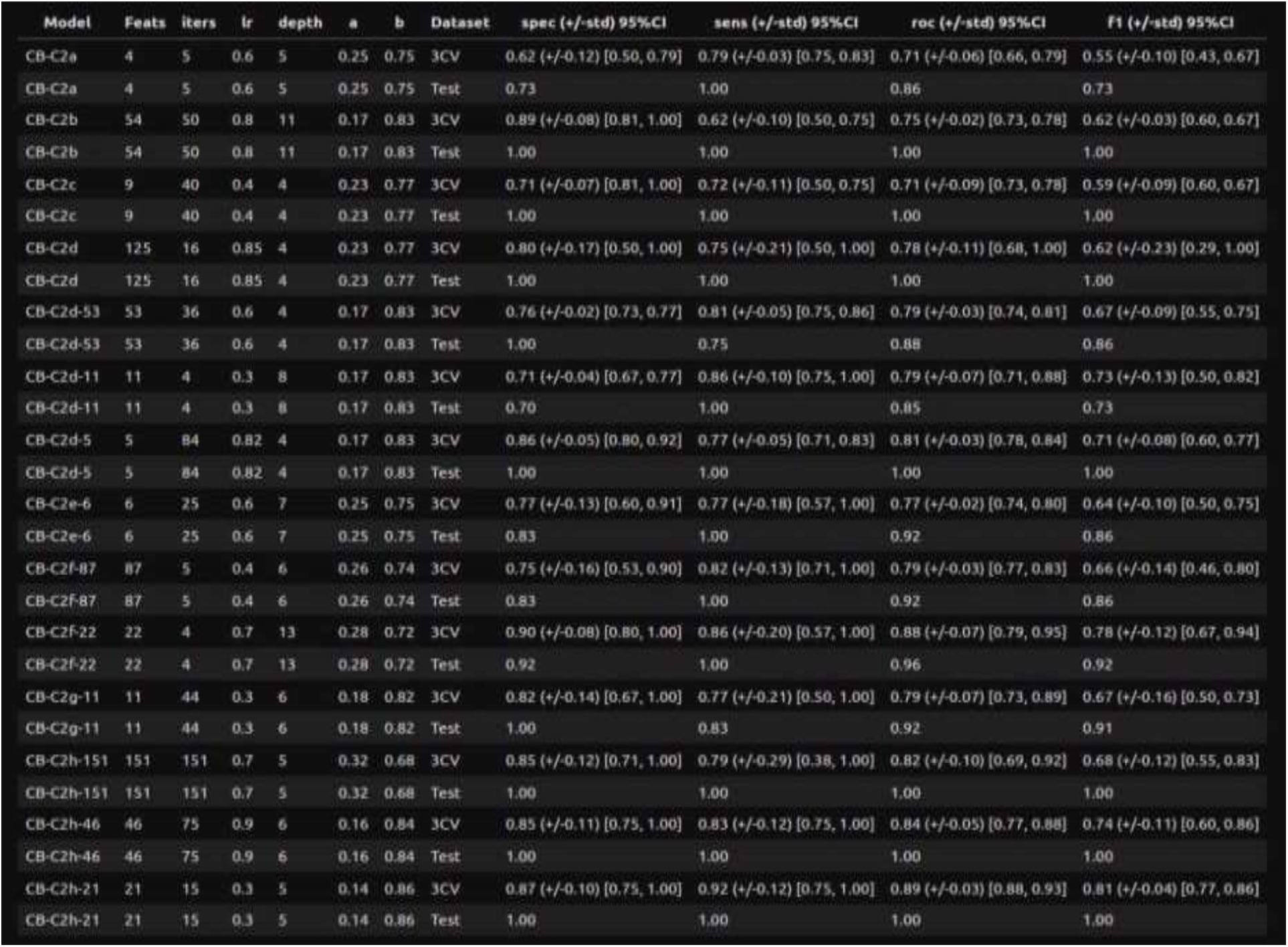
ML models of the C2: the name of the model, the number of features for each case, the values of their tuned hyper-parameters, the classes’ weights, the specificity, sensitivity, ROC-AUC, F1 scores, their mean and standard deviation as well as the 95% confidence interval for the 5CV on the train set and on the test set.

Correlation analysis showed that features like >150, >150 nm particle counts, BMI transformations, and weight-related interactions show some associations, but, similar to C1, none exceed the 0.6 threshold. This highlights that while certain features are associated with advanced steatosis, the strength of these associations are not even moderate, emphasizing the multifactorial nature of MASLD.

The plots in Figure 4 correspond to the C2 case, where S0, S1, and S2 are compared against S3. The violin plots (top panel) illustrate the distribution of various features across the two groups. Similar to the C1 case, the violin plots do not show a clear separation between the groups for any of the features, indicating that traditional visualization methods fail to provide significant insights or distinguishable patterns. In contrast, the SHAP scatter plots (bottom panel) reveal complex, non-linear relationships between features and the target variable. These plots highlight how features such as >150, >150 nm particle count, transformed weights, and BMI interactions contribute to the predictive model. The nuanced and dispersed patterns observed in the SHAP scatter plots underscore the importance of these features in classifying advanced steatosis (S3), despite the lack of clarity from traditional statistical methods.

**Figure 4.**
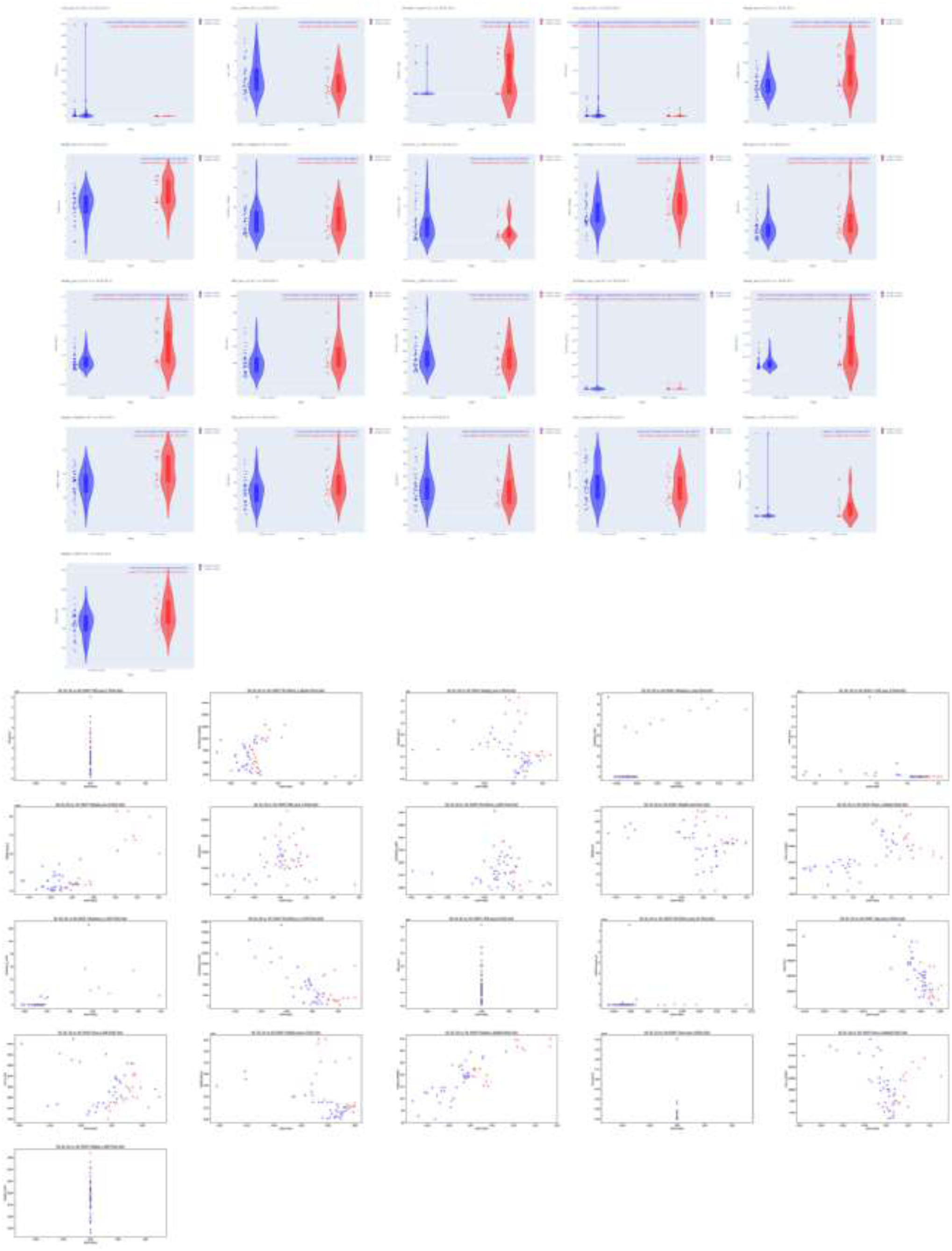
Plots of features distribution above and SHAP scatter plots for the train set of the ML CB-C2h-21 model.

In contrast to C1, the inclusion of anthropomorphic and clinical features, such as advanced fibrosis and diabetes, significantly enhanced model performance, increasing the AUROC from 0.81 to 0.89. The S3 group had a higher prevalence of advanced fibrosis (3/19), diabetes (9/19), and their co-occurrence (3/19) (Table 2), highlighting their relevance in identifying severe steatosis. Feature importance and SHAP analyses, shown in Figure 2 and Figure 4, revealed that EV features remained imporant predictors. Additionally, engineered features such as "Mean × Weight" and "Sum × Age" contributed significantly, indicating non-linear relationships between predictors and outcomes.

The consistent significance of EV across both cases underscores their diagnostic value. In C1, features like "Mean" and "50-150nm" were the strongest predictors, while in C2, engineered features such as "Mean × Weight" and "Sum_sqrt" played crucial roles. SHAP scatter plots in both cases (Figures 3 and 4) illustrated the absence of linear correlations, emphasizing the necessity of non-linear ML models to capture intricate interactions.

The XAI analysis provided interpretable insights into feature contributions. Figure 1 and Figure 3 highlighted "Mean" and "50-150nm" as the top predictors in C1, while Figure 2 and Figure 4 identified "Mean × Weight" and "Weight^7" as the most critical features in C2. These findings reinforce the physiological relevance of EV as biomarkers and their potential in MASLD diagnosis.

This study demonstrated the crucial role of EV in diagnosing and staging steatosis in MASLD. The CB-C1a model, which relied solely on EV, was sufficient for distinguishing S0 from S1, S2, and S3 stages in C1. Conversely, the CB-C2h-21 model showed that combining EV with anthropomorphic and clinical features improved the prediction of severe steatosis in C2. These results highlight the diagnostic potential of EV-related features and the utility of XAI in interpreting complex ML models for clinical applications in Hepatology.

## 4. Discussion

There is a continuous effort to identify new non-invasive biomarkers and tests for MASLD patients, due to the various limitations of liver biopsy. The development of blood biomarkers that can be reproducible, more easily applicable, and potentially available for utilization in clinical practice and treatment response monitoring, is in the spotlight of scientific research. Steatosis has a key role in the progression of the disease from MASL to MASH and fibrosis. Even though fibrosis strongly predicts long-term prognosis for MASLD patients and end-stage liver outcomes, early identification of steatosis can significantly alter the management of MASLD and its outcomes at hepatic and extrahepatic levels. In addition, there are notably fewer tests and biomarkers in the context of steatosis evaluation than in fibrosis, a phenomenon that implies an open horizon for new diagnostic perspectives.

Some of the limitation that have been reported regarding current biomarkers is the inclusion of age as a parameter for their calculation. More particularly, age as a parameter can importantly alter the interpretation of the results, a phenomenon that is mainly correlated with the significantly higher frequency of several diseases such as hyperlipidemia (increased triglyceride and cholesterol levels), insulin resistance, and diabetes (altered fasting glucose levels) due to aging [43].

Aging can also be implicated in the distribution of visceral fat, as well as in the noteworthy reduction of muscle mass, resulting in a normal BMI, altering the interpretation of several tests that include BMI [44]. Similarly, several enzymes can be influenced by aging and medication for age-related comorbidities, such as transaminases and GGT that may lead to inaccurate conclusions regarding steatosis [45]. Interestingly, a normal BMI can be also presented in lean patients with MASLD, which can lead to misinterpretation of the test, despite the presence of steatosis [46]. Similarly, some other anthropometric measurements such as waist-to-hip ratio or waist circumference also be normal in lean MASLD patients, leading to underestimated scores [47]. Meanwhile, it is reported that lean MASLD patients have a lower prevalence of hypertension, dyslipidemia, and T2DM, as well as lower levels of transaminases, higher HDL, and lower levels of TG and total cholesterol [47].

The absence of correlations exceeding 0.6 in both C1 and C2 suggests that no single feature serves as a strong predictor of steatosis stages in MASLD. This is consistent with the complex and multifactorial nature of the disease, where individual features may only have a limited explanatory or predictive role. The results emphasized the need for integrative approaches that combine multiple moderately correlated features to capture the heterogeneity of MASLD. The Pearson test identifies linear associations, while the Kendall and Spearman tests assess rank-based relationships, making them robust to non-linearity. The Point-Biserial test, tailored for binary classification, complements these analyses by quantifying the ability of features to separate distinct steatosis stages. Across all these methods, the moderate correlations (none exceeding 0.6) reinforce the idea that MASLD severity cannot be effectively characterized by isolated markers. This finding underlines the importance of ML models that leverage the combined contributions of several moderately associated features. Additionally, the moderate correlations align with clinical experience, where MASLD progression is driven by the interplay of metabolic, genetic, and environmental factors rather than a single dominant feature. These insights, derived from a robust and multi-faceted correlation analysis, provided a strong foundation for predictive modeling and biomarker discovery using ML and XAI.

The present study demonstrates the significant potential of EV as biomarkers for distinguishing steatosis stages in MASLD. By employing XAI and ML we elucidated the diagnostic importance of EV, particularly their size distributions and concentrations. The integration of advanced ML models like CB-C1a and CB-C2h-21 provides a robust framework for non-invasive, accurate diagnostics.

Figure 3 highlights the limitations of traditional statistical analysis and visualization techniques, such as violin plots, in capturing complex relationships within biological data, as demonstrated by the indistinguishable patterns that fail to show strong, separable linear relationships with steatosis stages in MASLD. In contrast, the SHAP scatter plots illustrate the power of machine learning and explainable AI (XAI) to detect and interpret non-linear associations, uncovering intricate patterns and feature interactions that are critical for understanding and differentiating steatosis stages, particularly in the C2 case where advanced steatosis (S3) is compared to earlier stages. The non-linear relationships revealed by SHAP emphasize the need for advanced analytical approaches to disentangle the multifactorial nature of MASLD and highlight the importance of features that might otherwise be overlooked due to their lack of linear separability. This ability to extract and interpret complex patterns not only enhances our understanding of MASLD but also compensates for the limitations of conventional methods, setting the stage for developing robust, data-driven predictive models. These findings demonstrate the transformative potential of XAI in hepatology research, paving the way for improved disease stratification and targeted interventions.

Our results underscore the central role of EV-related features in differentiating between various stages of steatosis. For C1 EV independently achieved high diagnostic performance, with the CB-C1a model achieving an AUROC of 0.70 in 5CV and 0.86 on the test set. Anthropomorphic and clinical features such as diabetes and advanced fibrosis did not improve diagnostic accuracy in this scenario. These findings highlight the potential of EV as standalone non-invasive biomarkers for early-stage steatosis detection. Conversely, for C2 the inclusion of anthropomorphic and clinical features significantly enhanced the model’s diagnostic performance, as evidenced by the CB-C2h-21 model’s AUROC improvement from 0.81 to 0.89 in 3CV. The higher prevalence of advanced fibrosis (3/19), diabetes (9/19), and their co-occurrence (3/19) in the S3 group suggests that these features are more relevant for identifying advanced stages of steatosis. Importantly, EV remained pivotal predictors in this context, underscoring their physiological relevance in MASLD progression. The results using 10 times iterative cross validation of C1 and C2 cases with AUROC +/-std of 0.71+/-0.03 and 0.81+/-0.04, respectfully, as well as the performance regarding the specificity, sensitivity and F1 score in Table 5, shows that even though the dataset volume was relatively low, the performance of the two best models was very good and promising. From the explainability and the insights on the associations of the most important features it is clear that VEs are potentially very strongly non-linear correlated with steatosis stages.

**Table 5.**
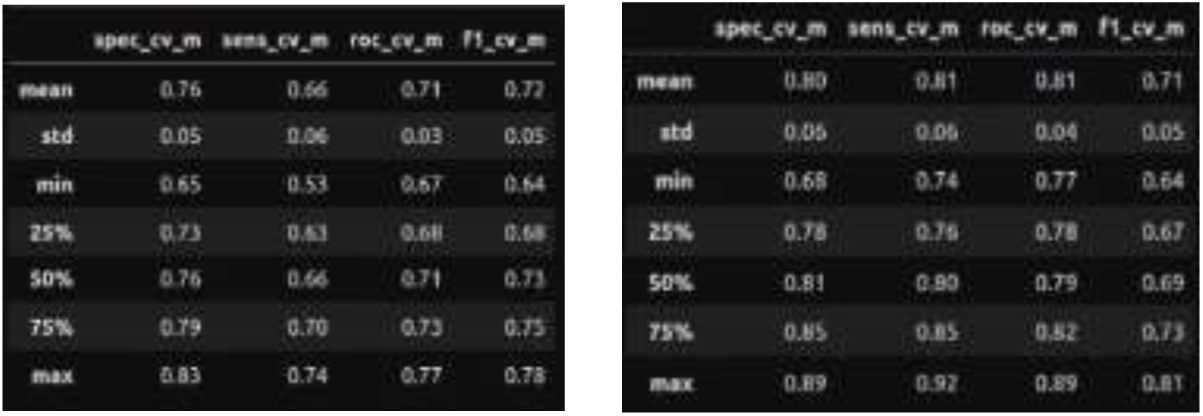
Performance on the the CB-C1a (left) and CB-C2h-21 (right) using 10 times iterative 5CV and 3CV, for C1 and C2 accordingly. Here x_cv_m for x in {spec, sens, roc, f1} corresponds to {specificity, sensitivity, AUROC, F1}.

The observed diagnostic performance aligns with the established role of EV in MASLD pathogenesis. EV, particularly those with specific size distributions, are known mediators of intercellular communication, inflammation, and angiogenesis. These mechanisms are crucial in the progression of MASLD from simple steatosis to advanced fibrosis and hepatocellular carcinoma. Our findings suggest that EV size distributions ("Mean," "50-150nm," ">150nm") and concentrations ("Sum") could serve as reliable, non-invasive biomarkers for disease staging and risk stratification.

By leveraging XAI tools such as SHAP analysis, we provided interpretable insights into the non-linear relationships between EV, anthropomorphic, clinical features, and steatosis stages. This transparency is critical for clinical adoption, as it enables healthcare professionals to understand the factors driving model predictions. The high sensitivity, specificity, and AUROC values achieved by CB-C1a and CB-C2h-21 models demonstrate their robustness and potentially clinical applicability.

Additionally, the use of a standardized protocol for EV analysis and rigorous validation of ML models ensures the reliability and reproducibility of our results. While liver biopsy remains the gold standard for steatosis assessment, our findings support the potential of ML-enhanced EV as non-invasive alternatives. Our approach could reduce the reliance on invasive procedures, making steatosis staging more accessible and patient-friendly.

Despite the promising findings, the study has some limitations such as the variability in blood sample processing and EV analysis that could possibly introduce bias [48–50]. More particularly, the count of particles could be influenced and overestimated due to similar-sized contaminants or lipoproteins [49,50], while serial centrifugations and freeze-thawing cycles can cause EV breakage [51]. Finally, another limitation of this study is the lack of biopsy validation for many of the patients, due to the ethical reasons, incompliance, and high cost of the procedure. However, we followed strict selection-criteria after excluding other causes of steatosis, other chronic liver diseases, and several conditions that can significantly alter the total circulating levels and the size of EVs.

Future research should explore the in-depth characterization of EV cargoes and their specific roles in MASLD progression, as well as the Apoptotic EVs (ApoEVs) that exceed 1000nm and their role in inflammation-related condition, such as lipotoxicity, their implication in tissue regeneration and their possible utilization as drug vectors [52]. Combining EV with advanced imaging techniques and other non-invasive biomarkers could further improve diagnostic accuracy.

## 5. Conclusion

This study demonstrates the potential of extracellular vesicles (EVs) as non-invasive biomarkers for diagnosing and staging steatosis in MASLD patients. Using advanced machine learning (ML) techniques combined with explainable artificial intelligence (XAI), we successfully identified EV, particularly their size distributions and concentrations, as critical features for differentiating between steatosis stages. The CB-C1a model achieved strong performance for distinguishing S0 from S1-S3 steatosis stages using EV alone, while the CB-C2h-21 model demonstrated the added diagnostic value of incorporating anthropomorphic and clinical features, such as diabetes and advanced fibrosis, for identifying severe steatosis (S3).

Correlation analysis revealed that no single feature exceeded a moderate threshold (0.6), emphasizing the multifactorial and non-linear nature of MASLD. Traditional statistical and visualization techniques, such as violin plots, failed to provide meaningful separations between groups, further highlighting the need for advanced analytical approaches. SHAP-based XAI enabled the detection of complex non-linear patterns, offering interpretable insights into feature contributions and their interactions, thus addressing the limitations of conventional methods.

These findings underscore the diagnostic utility of EVs in MASLD and the transformative potential of ML-enhanced EV analysis to provide scalable, patient-friendly alternatives to invasive liver biopsies. Future work should focus on further characterizing EV cargoes and integrating these biomarkers with other non-invasive diagnostic tools to refine disease staging and improve clinical outcomes in MASLD management.

## 6. Abbreviations

● Blood Processing Interval (BPI)
● Cross-Validation (CV)
● Data Science (DS)
● Explainable Artificial Intelligence (XAI)
● Extracellular Vesicles (EVs)
● Feature Engineering (FE)
● Feature Selection (FS)
● Interquartile Range (IQR)
● Liver Stiffness Measurement (LSM)
● Machine Learning (ML)
● Metabolic dysfunction-associated steatohepatitis (MASH)
● Metabolic dysfunction-associated steatotic liver disease (MASLD)
● Nanoparticle Tracking Analysis (NTA)
● Non-alcoholic Fatty liver disease (NAFLD)
● Non-invasive tests (NITs)
● Machine Learning Enhanced Non-invasive Tests (MLE-NITs)
● Steatotic Liver Disease (SLD).
● Type 2 Diabetes Mellitus (T2DM)
● Ultrasound Attenuation Parameter (UAP)
● Vibration-Controlled Transient Elastography (VCTE)

## Data Availability

All data produced in the present study are available upon reasonable request to the authors

---

**Figure 5.**
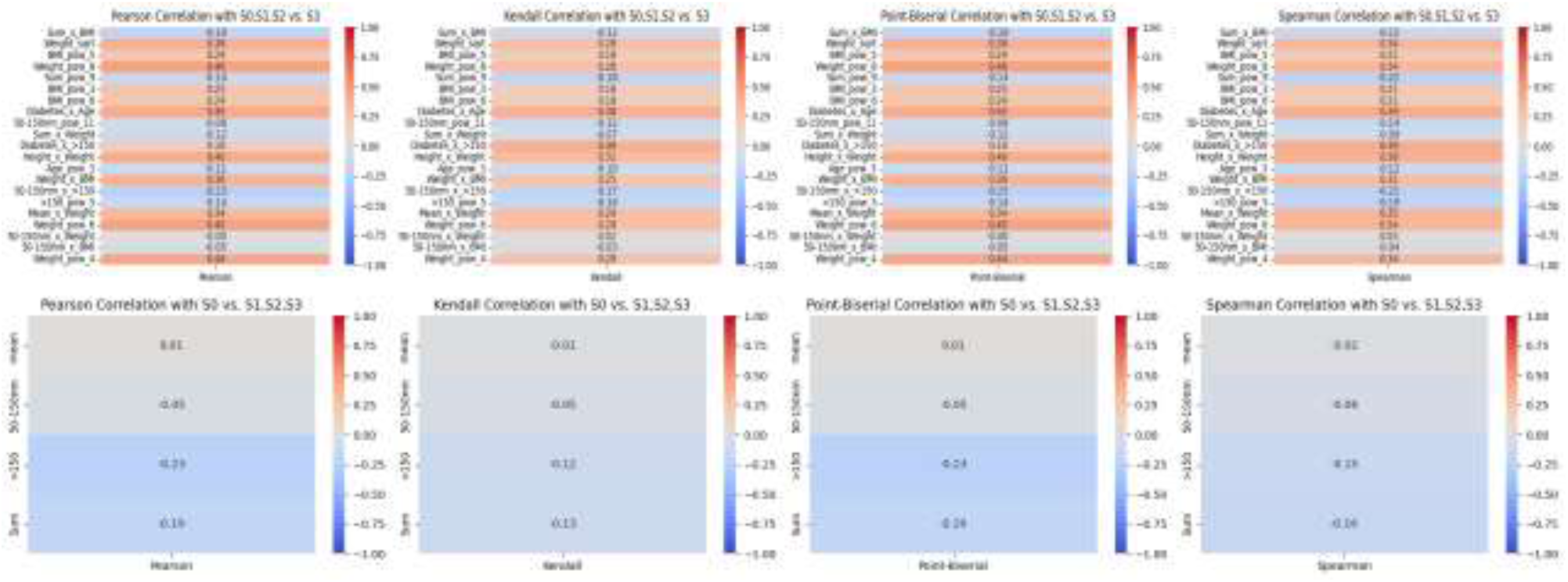
Plots of correlations of parametric and non-parametric tests for C1 and C2.

## Notes

### Competing Interest Statement

The authors have declared no competing interest.

### Funding Statement

This study did not receive any funding

### Author Declarations

Ethics Committee of General Hospital of Athens "Hippocratio" in 1st Health Authority of Greece ( Attica) gave ethical approval for this work

